# Dupilumab use is associated with protection from COVID-19 mortality: A retrospective analysis

**DOI:** 10.1101/2022.03.30.22273174

**Authors:** A. Donlan, I. Mallawaarachchi, J. Sasson, R. Preissner, J. Loomba, W.A. Petri

## Abstract

We previously found that type 2 immunity promotes COVID-19 pathogenesis in a mouse model. To test relevance to human disease we used electronic health record databases and determined that patients on dupilumab (anti-IL-4R*α* monoclonal antibody that blocks IL-13 and IL-4 signaling) at the time of COVID-19 infection had lower mortality.

## Introduction

Severe Acute Respiratory Syndrome Coronavirus 2 (SARS-CoV-2) is the virus that causes Coronavirus Disease 2019 (COVID-19) and is currently causing a devastating global pandemic. Many approaches to combat mortality involve targeting inflammation, such as with the corticosteroid dexamethasone (The RECOVERY Collaborative Group 2021) or the monoclonal antibody against IL-6, tocilizumab (Abani et al. 2021). However, while these therapeutic options have been observed to reduce mortality in patients, protection is not complete. This was exemplified by mortality due to the delta variant being 12.6% during the fall 2021 surge (Iuliano et al. 202AD). More recently, in hospital deaths due to the omicron variant dropped to 7.3% (Iuliano et al. 202AD), however, reported deaths still remain high and methods to reduce these numbers are warranted. Additionally, the emergence of novel variants, such as omicron, for which the vaccine is increasingly less effective against highlights that we need ongoing development and identification of better therapeutic options in treating this disease for those who become ill.

Recently, we have uncovered a causal role of Interleukin (IL)-13 in promoting severe outcomes caused by infection with SARS-CoV-2, suggesting that type 2 immune responses are pathogenic during COVID-19 (Donlan et al. 2021). IL-13 is often associated with promoting pathology in asthma, allergies, and atopic dermatitis. In the lung, pulmonary responses potentiated by IL-13 include airway hyperreactivity, mucus production, smooth muscle contractility, recruitment of immune cells, and long-term airway remodeling (Seibold 2018; Wills-Karp et al. 1998; Zhu et al. 1999). In acute settings this results in airway restriction causing breathing difficulty and wheezing, and long-term can result in decreased lung capacity and function.

Dupilumab is a human monoclonal antibody which blocks signaling of the closely related cytokines IL-4 and IL-13 by targeting the shared alpha subunit of their receptors, IL-4R*α* (Wenzel et al. 2013; Beck et al. 2014; Castro et al. 2018). IL-4 and IL-13 are both primarily associated with type 2 responses which drive pathogenesis of asthma and atopic dermatitis, for which dupilumab is approved to treat (Wenzel et al. 2013; Beck et al. 2014; Castro et al. 2018). We were interested in whether dupilumab use in patients who were later diagnosed with COVID-19 was associated with protection from mortality due to its ability to block pathogenic IL-13 signaling. Earlier we had found in a small number (81 total) of patients an association of dupilumab use with protection from death due to COVID-19 (Donlan et al. 2021).

Here we report for over a thousand patients that dupilumab usage is associated with reduced mortality compared to matched-control patients. Our findings support the hypothesis that IL-13 signaling during COVID-19 is associated with more severe outcomes, and that pharmacological inhibition of this pathway may be a feasible therapeutic option for treating this disease.

## Methods

The N3C data transfer to NCATS is performed under a John Hopkins University Reliance Protocol # IRB00249128 or individual site agreements with NIH. The N3C Data Enclave is managed under the authority of the NIH; information can be found at https://ncats.nih.gov/n3c/resources.

### Database and Inclusion Criteria

#### N3C

The N3C (National COVID Cohort Collaborative) cohort was filtered to patients who were COVID-19 positive based on the presence of either a SARS-CoV-2 PCR or antigen-positive test results or a COVID-19 diagnosis code. The first instance of either indicator in the patient’s record was used as the COVID-19 index date. To account for dupilumab use in COVID-19 patients that would be far removed from their diagnosis, and therefore unlikely to be exerting any pharmacological effects, a cutoff was made for dupilumab use within 61 days prior to the patients’ index event. Using these filters, there were three sub cohorts that were analyzed:

1. Controls: COVID-19 positive patients with no record of dupilumab use within 61 days of their index event.
2. Dupilumab (+): Any COVID-19 patient with recorded dupilumab use within the 61 days preceding their index event.
3. Dupilumab (-): COVID-19 negative patients with recorded dupilumab use.

The incidence of COVID positivity in people on dupilumab [cohort definition 1 above / (definition 1 + 2)], along with 95% confidence intervals, was calculated.

#### TriNetX

Data were retrieved from the COVID-19 Research Network provided by TriNetX, comprising 80 million patients from 65 health care organizations in 11 countries (for database access on 12/07/2021). COVID-19 patients were identified via the ICD-10 code U07.1 or the presence of a SARS-CoV-2-related RNA diagnosis within the last two years. Drug use was identified via RxNorm codes for dupilumab (1876376) and the lab value for C-reactive protein (9063). 1:1 matching was performed for age and sex.

### Matching

#### N3C

A case-control design was used. Dupilumab (+) patients were matched to control patients in a 1:5 ratio, with exact matching on gender, race and ethnicity, N3C site, asthma and nearest matching on age (**Supplemental Table 1**).

#### TriNetX

Analytical tools were used to obtain baseline characteristics, balance cohorts with propensity score matching and analyze outcomes of interest in the final cohorts. Baseline characteristics, including demographics, diagnoses, procedures, and medication were obtained. Propensity score matching was used to balance cohorts. Propensity scores matched cohorts 1:1 using a nearest neighbor greedy matching algorithm with a caliper of 0.25 times the standard deviation.

### Outcome Definitions

1. Hospitalized: The COVID-19 (+) test or diagnosis code occurred during a hospitalization (any consecutive series of visit encounters that include an inpatient stay).
2. Death: Death as recorded in medical record system from contributing sites.

COVID Severity: Categorical variable with following levels:

1. Mild - Outpatient
2. Moderate - Hospitalized but no ECMO or IMV
3. Severe - Hospitalized with ECMO or IMV
4. Death

### Statistical Analysis

#### N3C

Statistical analyses were performed using R studio version 3.5.1. Conditional logistic regression, or exact tests in rare outcomes, was used to compare COVID-19 severity outcomes within the matched subset of COVID (+) patients.

#### TriNetX

Outcomes were defined as ventilation assist and death. Measures of association including risk differences with their respective 95% CI’s were calculated. Time frame of follow-up for both groups was set to 365 days for Kaplan-Meier curves, which were generated for each analysis.

## Results

The N3C and TriNetX databases were independently queried. The N3C database as of August 27^th^ 2021 included 1069 patients prescribed dupilumab for which 220 were subsequently diagnosed with COVID-19 within 61 days of their dupilumab dose (infection rate 20.6%; 95% CI: 18.2%-23.1%). We found that dupilumab use was associated with significantly fewer deaths than in the matched control group (0 vs 24 (2.2%); OR: 0.02) (**Table 1**).

**Table 1.** Disease outcomes in patients taking dupilumab compared to matched controls.

| **A. N3C** |  |  |  |  |
| --- | --- | --- | --- | --- |
|  | **Controls (N=1100)** | **Dupilumab (N=220)** | **Odds Ratio** | **P-value** |
| **Outcome** |  |  |  |  |
| Hospitalized | 111 (10.1%) | 22 (10.0%) | 0.99 | 0.97 |
| Died | 24 (2.2%) | 0 (0.0%) | 0.02 | <.001 |
| **COVID-19 Severity** |  |  |  |  |
| Mild | 982 (89.3%) | 198 (90.0%) |  |  |
| Moderate | 83 (7.5%) | 20 (9.1%) |  |  |
| Severe | <20** | <20** |  |  |
| Death | 24 (2.2%) | 0 (0.0%) |  |  |
| **B. TriNetX** |  |  |  |  |
|  | **Controls (N=1085)** | **Dupilumab (N=1085)** |  | **Log-rank p-value** |
| Died | 15* | 10* |  |  |
| Survival Probability | 96.91% | 98.84% |  | 0.03 |

\*\*; To protect patient privacy, suppressed in accordance with N3C download policy
\*; These data are derived directly from Trinetx and therefore not subject to N3C download policy.

Next, to support the N3C findings, we utilized the TriNetX database and filtered for COVID-19 cases with (n= 937) or without (n= 1.7 million) recorded use of dupilumab and performed 1:1 matching (**Supplemental Table 2**). We found that dupilumab usage was associated with a lower risk of death (log-rank p-value = 0.03) (**Table 1**) for death when compared to controls. Additionally, dupilumab usage in this cohort was associated with decreased CRP values, indicating that inflammation was likely lower in this group.

## Discussion

Through utilization of two databases, we have found that prior dupilumab usage in COVID-19 patients was associated with improved survival compared to matched controls. Previous work by Ungar et al., 2022 supported this finding, where, in atopic dermatitis patients dupilumab use was also associated with a reduction in severe outcomes from COVID-19 (Ungar et al. 2022). We report these findings building off of *in vivo* work supporting a causal role for IL-13 in COVID-19 pathogenesis. Due to the mechanism of action of dupilumab, we hypothesize this protection, then, is mediated by blocking pathogenic IL-13 signaling.

Retrospective analyses, such as these, provide us with large-scale data that allow for smaller confidence intervals than smaller prospective studies. However, there are limitations due to non-randomized groups resulting in sampling biases, difficulty defining temporal boundaries, and not being able to infer causational relationships. The N3C database used allowed for well-defined patient outcomes and temporal windows, however small sampling size may limit the statistical power through this method. Supportive analysis by TriNetX allowed for larger cohort of dupilumab (+) cases, but was limited to lower matching criteria and at a 1:1 ratio. Utilization of both datasets, then, provides two analyses which supported our hypothesis.

Identification of dupilumab as a being associated with reduction in death due to COVID-19 may implicate this drug as a potential therapeutic option for patients. Future, large-scale clinical trial of dupilumab use during COVID-19 will be important for understanding the impact this drug may have on protecting patients from severe outcomes.

Author contributions: AD wrote the manuscript. AD, IM, JL, and RB assisted with data analysis. All authors contributed to discussion regarding conceptualization and design of the reported studies. Authorship was determined using ICMJE recommendations.

### Attribution

This research was possible because of the patients whose information is included within the data and the organizations and scientists who have contributed to the on-going development of this community resource https://doi.org/10.1093/jamia/ocaa196.

## Data Availability

All data produced in the present study are available upon reasonable request to the authors

## Funding Sources

AD, JS, and WP were supported by grants to WP from the Manning Family Foundation, Ivy Foundation, Henske Family, and NIH R01 AI124214.

Analyses performed by IM and JL were conducted with data or tools accessed through the NCATS N3C Data Enclave https://covid.cd2h.org and N3C Attribution & Publication Policy v1.2-2020-08-25b, and supported by NCATS U24 TR002306. Support for project planning and analysis was also provided by the integrated Translational Health Research Institute (iTHRIV) with funding support from NCATS UL1 TR003015. RP was supported by Deutsche Forschungsgemeinschaft (DFG KFO339, DFG TRR295).

## Competing interests

William A. Petri, Jr. receives research funding from Regeneron, Inc. which is the maker of dupilumab. The other authors declare no competing interests.

## Tables

Table S1. Patient demographics in N3C cohort.

Table S2. Patient demographics in TriNetX cohort.

Table S3. Dupilumab use is associated with lower CRP levels of patients in TriNetX database.

## Notes

### Author Declarations

N3C Publication Committee gave ethical approval for this work.

## References

Abani, Obbina, Ali Abbas, Fatima Abbas, Mustafa Abbas, Sadia Abbasi, Hakam Abbass, Alfie Abbott, et al. 2021. “Tocilizumab in Patients Admitted to Hospital with COVID-19 (RECOVERY): A Randomised, Controlled, Open-Label, Platform Trial.” The Lancet 397 (10285): 1637–45. https://doi.org/10.1016/S0140-6736(21)00676-0.

Beck, Lisa A., Diamant Thaçi, Jennifer D. Hamilton, Neil M. Graham, Thomas Bieber, Ross Rocklin, Jeffrey E. Ming, et al. 2014. “Dupilumab Treatment in Adults with Moderate-to-Severe Atopic Dermatitis.” New England Journal of Medicine 371 (2): 130–39. https://doi.org/10.1056/NEJMoa1314768.

Castro, Mario, Jonathan Corren, Ian D. Pavord, Jorge Maspero, Sally Wenzel, Klaus F. Rabe, William W. Busse, et al. 2018. “Dupilumab Efficacy and Safety in Moderate-to-Severe Uncontrolled Asthma.” New England Journal of Medicine 378 (26): 2486–96. https://doi.org/10.1056/NEJMoa1804092.

Donlan, Alexandra N., Tara E. Sutherland, Chelsea Marie, Saskia Preissner, Benjamin T. Bradley, Rebecca M. Carpenter, Jeffrey M. Sturek, et al. 2021. “IL-13 Is a Driver of COVID-19 Severity.” JCI Insight, June. https://doi.org/10.1172/jci.insight.150107.

Iuliano, A. Danielle, Joan M. Brunkard, Tegan K. Boehmer, Elisha Peterson, Stacey Adjei, Alison M. Binder, Stacy Cobb, et al. 202AD. “Trends in Disease Severity and Health Care Utilization During the Early Omicron Variant Period Compared with Previous SARS-CoV-2 High Transmission Periods — United States, December 2020–January 2022.” CDC. https://www.cdc.gov/mmwr/volumes/71/wr/mm7104e4.htm#suggestedcitation.

Seibold, Max A. 2018. “Interleukin-13 Stimulation Reveals the Cellular and Functional Plasticity of the Airway Epithelium.” Annals of the American Thoracic Society 15 (Supplement_2): S98–102. https://doi.org/10.1513/AnnalsATS.201711-868MG.

The RECOVERY Collaborative Group. 2021. “Dexamethasone in Hospitalized Patients with Covid-19.” New England Journal of Medicine 384 (8): 693–704. https://doi.org/10.1056/NEJMoa2021436.

Ungar, Benjamin, Jacob W. Glickman, Alexandra K. Golant, Celina Dubin, Olga Marushchak, Alyssa Gontzes, Daniela Mikhaylov, et al. 2022. “COVID-19 Symptoms Are Attenuated in Moderate-to-Severe Atopic Dermatitis Patients Treated with Dupilumab.” The Journal of Allergy and Clinical Immunology: In Practice 10 (1): 134–42. https://doi.org/10.1016/j.jaip.2021.10.050.

Wenzel, Sally, Linda Ford, David Pearlman, Sheldon Spector, Lawrence Sher, Franck Skobieranda, Lin Wang, et al. 2013. “Dupilumab in Persistent Asthma with Elevated Eosinophil Levels.” New England Journal of Medicine 368 (26): 2455– 66. https://doi.org/10.1056/NEJMoa1304048.

Wills-Karp, M., J. Luyimbazi, X. Xu, B. Schofield, T. Y. Neben, C. L. Karp, and D. D. Donaldson. 1998. “Interleukin-13: Central Mediator of Allergic Asthma.” Science 282 (5397): 2258–61. https://doi.org/10.1126/science.282.5397.2258.

Zhu, Zhou, Robert J. Homer, Zhonde Wang, Qingsheng Chen, Gregory P. Geba, Jingming Wang, Yong Zhang, and Jack A. Elias. 1999. “Pulmonary Expression of Interleukin-13 Causes Inflammation, Mucus Hypersecretion, Subepithelial Fibrosis, Physiologic Abnormalities, and Eotaxin Production.” Journal of Clinical Investigation 103 (6): 779–88. https://doi.org/10.1172/JCI5909.

