## Supplemental Tables for "Dupilumab use is associated with protection from COVID-19 mortality: A retrospective analysis"

**Supplemental Table 1. N3C Cohort**

|  | **Controls (N=1100)** | **Dupilumab+ (N= 220)** |
| --- | --- | --- |
| **Continuous Variable: mean (SD)** |  |  |
| Age | 46.3 (21.0) | 46.3 (21.0) |
| **Categorical variables: n (%)** |  |  |
| Age Group |  |  |
| 05-40 | 417 (38.1%) | 83 (37.9%) |
| 41-85 | 678 (61.9%) | 136 (62.1%) |
| Gender: Male | 420 (38.2%) | 84 (38.2%) |
| Race/Ethnicity: |  |  |
| Asian Non-Hispanic | suppressed | suppressed |
| Black or African American Non-Hispanic | 200 (18.2%) | 40 (18.2%) |
| Hispanic or Latino any Race | 130 (11.8%) | 26 (11.8%) |
| Other Non-Hispanic | suppressed | suppressed |
| Unknown | 120 (10.9%) | 24 (10.9%) |
| White Non-Hispanic | 615 (55.9%) | 123 (55.9%) |
| Asthma:Yes | 585 (53.2%) | 117 (53.2%) |

**Supplemental Table 2. TriNetX Cohort**

|  | **Controls (N=937)** | **Dupilumab+ (N= 937)** |
| --- | --- | --- |
| **Continuous Variable: mean (SD)** |  |  |
| Age | 44.7 (18.6) | 44.7 (18.6) |
| **Categorical variables: n (%)** |  |  |
| Gender: Male | 422 (38.894%) | 422 (38.894%) |

**Supplemental Table 3. CRP Values**

| **Group** | **Mean CRP (mg/L)** | **Median CRP (mg/L)** | **SD** |
| --- | --- | --- | --- |
| Died | 85.2 | 49.1 | 93.8 |
| W/o Dupilumab | 34.6 | 10 | 58.2 |
| W/ Dupilumab | 22.7 | 7 | 47.2 |
